# COVID-19 and Inflammatory Bowel Diseases: risk assessment, shared molecular pathways and therapeutic challenges

**DOI:** 10.1101/2020.04.28.20082859

**Authors:** Iolanda Valentina Popa, Mircea Diculescu, Cătălina Mihai, Cristina Cijevschi-Prelipcean, Alexandru Burlacu

## Abstract

**Background:** The novel coronavirus SARS-CoV-2 causing COVID-19 disease is yielding a global outbreak with serious threats to public health. In this paper, we aimed to review the current knowledge about COVID-19 infectious risk status in inflammatory bowel disease (IBD) patients requiring immunosuppressive medication. Also, we focused on several molecular insights that could explain why IBD patients appear to not have higher risks of infection and worse outcome in COVID-19 than the general population, in attempt to provide scientific support for safer decisions in IBD patient care.

**Methods:** PubMed electronic database was interogated for relevant articles involving data about common molecular pathways and shared treatment strategies between SARS-CoV-2, SARS-CoV-1, MERS-CoV and inflammatory bowel diseases. In addition, Neural Covidex, an artificial intelligence tool, was used to answer queries about pathogenic coronaviruses and possible IBD interactions using the COVID-19 Open Research Dataset (CORD-19).

**Discussions:** Few molecular and therapeutic interactions between IBD and pathogenic coronaviruses were explored. First, we showed how the activity of soluble angiotensin-converting enzyme 2, CD209L alternate receptor and phosphorylated *α* subunit of eukaryotic translation initiation factor 2 might exert protective impact in IBD in case of coronavirus infection. Second, IBD medication was discussed in the context of possible beneficial effects on COVID-19 pathogeny including “cytokine storm” prevention and treatment, immunomodulation, interferon signaling blocking, viral endocytosis inhibition.

**Conclusions:** Using current understanding of SARS-CoV-2 as well as other pathogenic coronaviruses immunopathology, we showed why IBD patients should not be considered at an increased risk of infection or more severe outcomes. Whether our findings are entirely applicable to the pathogenesis, disease susceptibility and treatment management of SARS-CoV-2 infection in IBD must be further explored.

## 1. Introduction

The emergence of novel coronavirus SARS-CoV-2 that causes the COVID-19 disease is yielding a global outbreak with serious threats to public health due to a very high transmissibility rate, being spread in 213 countries with 1,569,504 confirmed COVID-19 cases and 95,269 confirmed deaths as of April 11^th^ 2020 [1] and due to potential severe complications in elderly or comorbid patients [2].

Given the high infectivity, the complexity of immune mechanisms involved in both SARS-CoV-2 infection as well as in inflammatory bowel diseases (IBD), the immunosuppresive therapy [3] and the gastrointestinal events reported in numerous COVID-19 patients [4], the problem of particular evolution and treatment management in infected IBD patients is of real concern. Although, IBD causes are not known, autoimmunity and immune-mediated mechanisms have an important role to play in disease pathogenesis [3] and immunosuppressive and immunomodulating drugs are successfully used in IBD therapy [5].

Main questions to answer are whether IBD patients are at an increased risk of SARSCoV-2 infection in the context of their immunosuppressive or immunomodulating treatment? Or do they have a higher risk of severe clinical course considering that their IBD diagnosis is a comorbidity? Should they stop or change current treatment knowing that this might expose them to develop an IBD flare? Might the two diseases share common molecular pathways that could influence each other’s evolution? Does IBD medication target COVID-19 pathways? How are current COVID-19 experimental therapies influence IBD course and relapses?

In this paper, we aimed to review the current knowledge about COVID-19 infectious risk status in IBD patients and to describe several molecular insights that could explain why IBD patients appear to not have higher risks of infection and worse outcome in COVID-19 than the general population, in the attempt to provide scientific support to current data for better, safer and more precise decisions in IBD patient care in epidemiological context.

## 2. Methods

The electronic database of PubMed was systematically searched for relevant articles from the inception until April 2020. The search terms used were [“*SARS-CoV-2*” OR “*COVID-19*” OR “*SARS*” OR “*SARS-CoV*” OR “*SARS-CoV-1*” OR "*MERS-CoV*"] AND [“*inflammatory bowel diseases*” OR “*ulcerative colitis*” OR “*Crohn*”] AND ["*molecular pathways*" OR "*immunosuppresive*"]. Study selection process included article identification, removing the duplicates, screening titles and abstracts, and assessing eligibility of the selected full texts. Additionally, reference lists of valid articles were checked for studies of relevance. Articles were included if they involved data about common molecular pathways and shared treatment strategies between SARS-CoV-2, SARS-CoV-1, MERS-CoV and IBD. Journal articles published with full text or abstracts in English were eligible for inclusion.

Additionally, the search engine https://covidex.ai made by the University of Waterloo and New York University was interogated. Neural Covidex uses natural language processing, state-of-the-art neural network models and artificial intelligence techniques to answer queries about pathogenic coronaviruses using the COVID-19 Open Research Dataset (CORD-19). CORD-19 is the current largest open dataset available with over 47000 scholarly articles, including over 36000 with full text about COVID-19, SARS-CoV-2 and other coronaviruses from the following sources: PubMed’s PMC open access corpus, a corpus maintained by the WHO, bioRxiv and medRxiv preprints. The CORD-19 dataset is available at https://pages.semanticscholar.org/coronavirus-research.

Neural Covidex was interogated with the following queries: “SARS-CoV-2 and inflammatory bowel diseases”, “COVID-19 and inflammatory bowel diseases”, “SARS and inflammatory bowel diseases”, “MERS-CoV and inflammatory bowel diseases”. The first interogation “SARS-CoV-2 and inflammatory bowel diseases” returned 40 results. Second query resulted in 25 articles. Third interogation returned 39 articles, and the fourth query lead to 31 results. After removing the duplicates and assessing the relevance for the research subject, 48 articles were included in the study.

Study selection process and number of papers identified in each phase are illustrated in the Flowchart (Figure 1).

**Figure 1 Legend.**
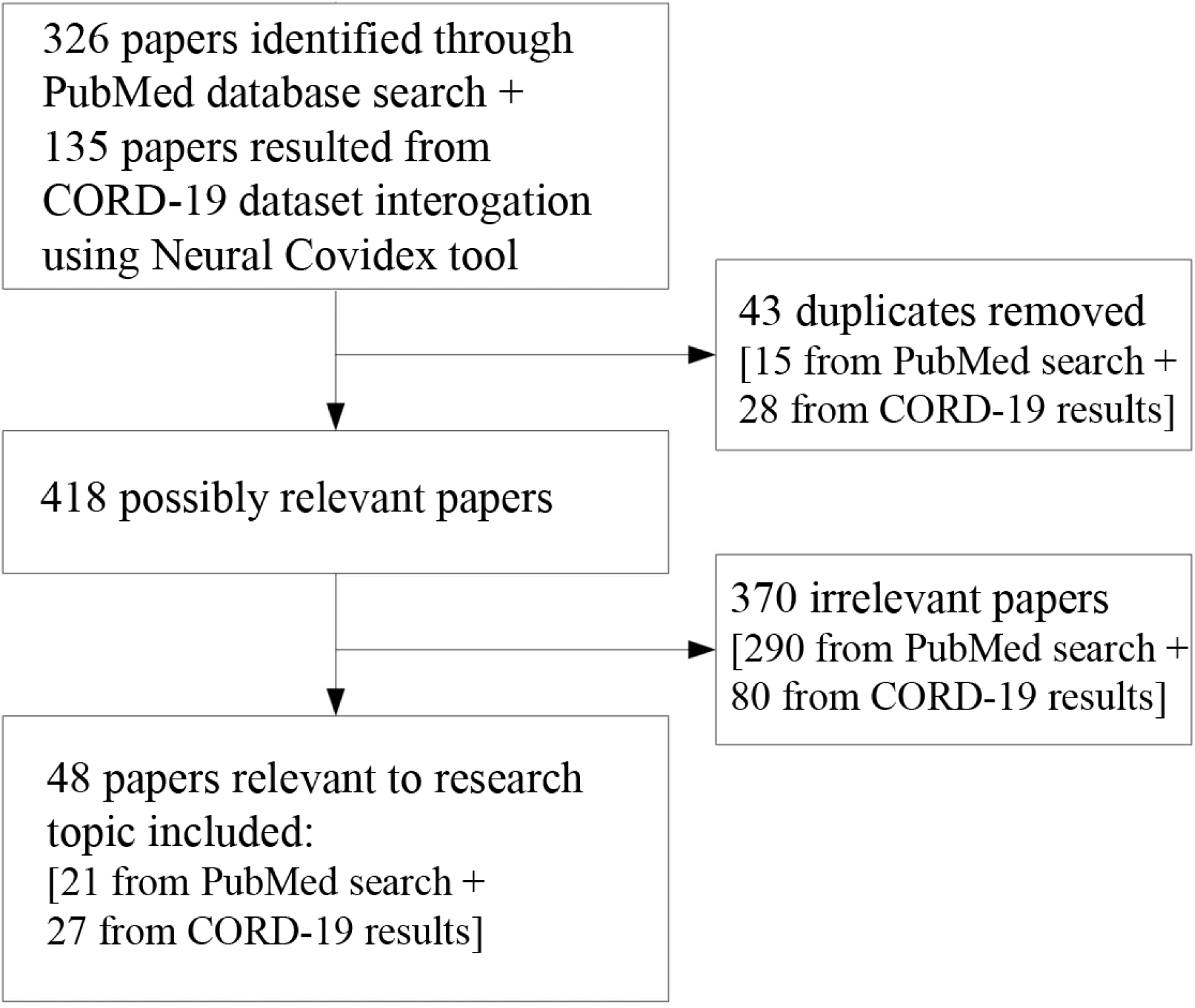
Study selection process and number of papers included.

## 3. Discussions

### a. Why IBD patients are expected to be more vulnerable than general population facing COVID-19 threat?

It has been shown that IBD patients are at increased risk of pneumonia [6], influenza infection [7] and other infectious complications compared to non-IBD population [8].

Current medication used in IBD consist of anti-inflammatory drugs (Mesalazine, corticosteroids), immunosuppresives (Azathioprine, 6-mercaptopurine, Methotrexat) and biologics (Infliximab and Adalimumab – anti-tumor necrosis factor antibodies, Vedolizumab – monoclonal antibody with gut selectivity that inhibits α_4_β_7_ integrin, Ustekinumab – IL-12 and IL-23 antagonist, Tocilizumab – anti-IL-6R antibody, Tofacitinib – Janus kinase inhibitor) [9]. IBD patients treated with azathioprine/6-mercaptopurine or immunosuppressant combination therapy and patients older than 50 have been shown to present an increased risk of opportunistic infections [10,11]. A meta-analysis of randomized control trials comparing anti-tumor necrosis factor (TNF) therapy with placebo found that anti-TNF therapy significantly increases the risk of opportunistic infections in IBD patients [12,13]. Vedolizumab has been linked to respiratory and bowel infections, although to a lesser extent than anti-TNF medication [14].

Moreover, recent studies described concomitant digestive symptoms (particularly diarrhea and abdominal pain) associated with COVID-19 [4,15]. Also, there has been a case report about SARS-CoV-2 gastrointestinal infection causing acute hemorrhagic colitis with colonic injury confirmed endoscopically for which other etiologies have been excluded [16].

In addition, pre-existing digestive diseases like hepatitis B infection and liver injury are more prevalent in severe COVID-19 cases than in mild ones [2]. Coronaviruses bind to their target cells through angiotensin-converting enzyme 2 (ACE2). Particularly high expressions of ACE2 can be found in ileum, colon and other digestive tract segments, testis, renal, cardiovascular and lung tissues [17]. Notably, studies reveal that the inflamed gut in IBD patients is associated with higher ACE2 expression [18] which may facilitate viral entrance.

Finally, coronavirus spike protein, that binds to ACE2 receptor, is the substrate for a trypsin-like proteinase found on host cell surface. Trypsin-like proteinase activates the viral spike protein, which in turn initiates fusioning process between the virus and the host cell. It has been shown that several fecal serine proteinases (including the trypsin-like protease) have increased activity in IBD [19,20].

### b. Real-world-data and current experts’ opinions in IBD and COVID-19

Even though expected consequences of IBD patient’s infection with the novel coronavirus are based on a solid reasoning, reality does not seem to confirm anticipated poor outcomes.

Currently, the only existing patient data are synthesized in SECURE-IBD, an international IBD reporting database, comprising of 457 patients infected with SARS-CoV-2 (as of April 11^th^ 2020, available at https:/covidibd.org/). This database offers information about number of cases by country and outcomes by patient characteristics (age, sex, diagnosis, disease activity, comorbidities, medication). Current data does not reveal higher risks of infection, more severe outcomes or significant differences in disease course between IBD patients and non-IBD patients infected with SARS-CoV-2.

According to SECURE-IBD database, 30% of patients with IBD and SARS-CoV-2 required hospitalization, 7% had a severe disease course either requiring intensive care, invasive ventilation or died, the death rate being 3% (Brenner EJ, Ungaro RC, Colombel JF, Kappelman MD. SECURE-IBD Database Public Data Update. covidibd.org. Accessed on 04/11/20, https://covidibd.org/current-data/).

As the American College of Gastroenterology (ACG) states, currently available information does not attest higher risk of SARS-CoV-2 infection or COVID-19 development for IBD patients, whether or not they are under treatment [21]. Also, at the 2020 meeting of the International Organization for the Study of Inflammatory Bowel Diseases, members and selected content experts voted the appropriateness of multiple statements related to COVID-19 risks. Thereby, current reasoning was that the risk of infection with SARS-CoV-2 was the same whether a patient had IBD or did not have IBD and that patients with IBD who had COVID-19 did not have a higher mortality compared to patients without IBD [22].

### c. IBD patients share similar risk of SARS-CoV-2 infection as general population: possible counterintuitive arguments

Known and possible interactions between IBD and SARS-CoV-2 immunopathology concerning certain molecular pathways and treatment dynamics are summarized in Figure 2.

**Figure 2 Legend.**
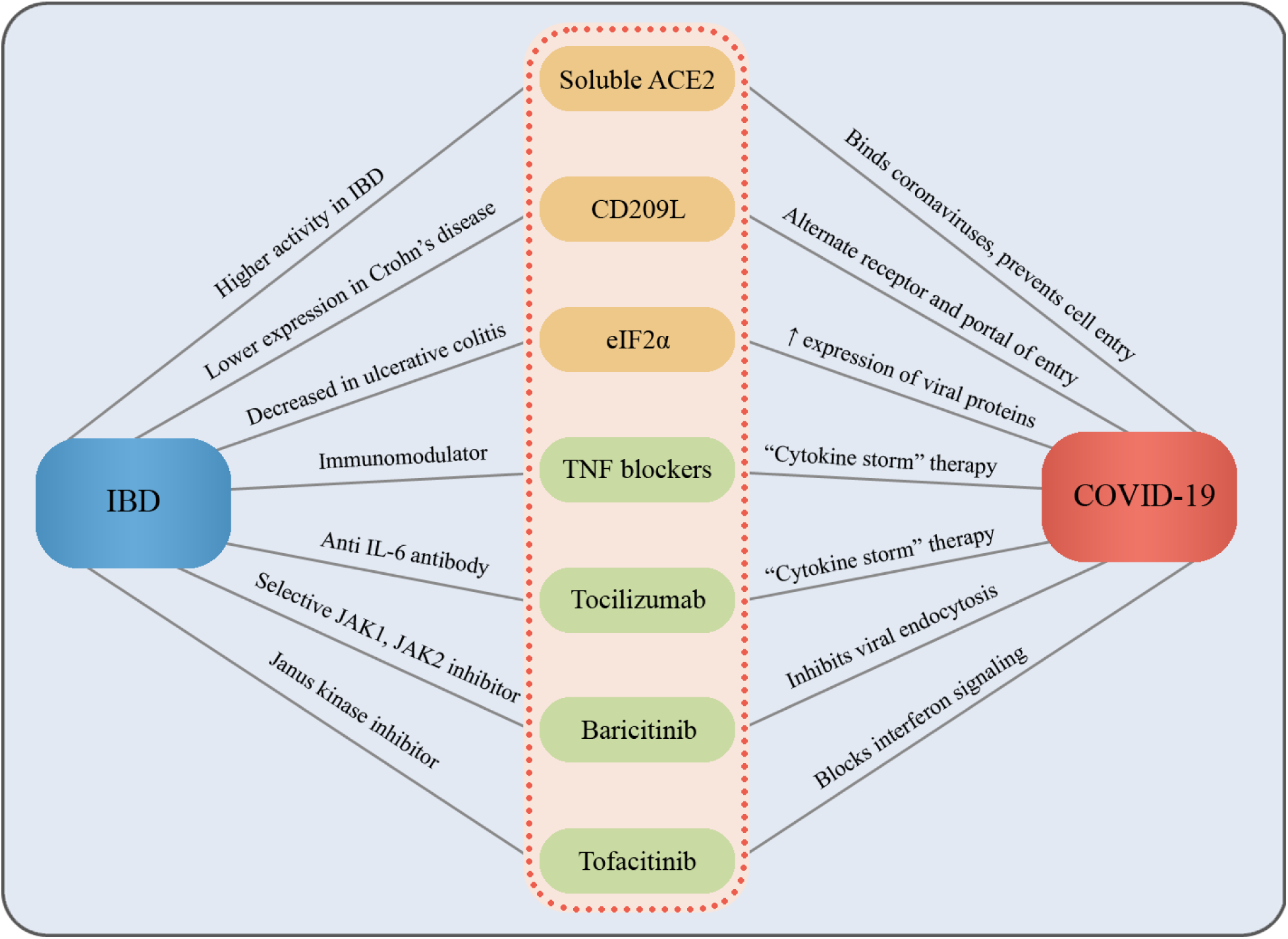
Known and possible interactions between IBD and SARS-CoV-2 immunopathology.

#### Molecular insights

Although, SARS-CoV-2 binds to the cells through ACE2 receptor that appears to have higher expression in IBD, one study found that ACE2 expression in the colonocytes was positively associated with genes regulating viral infection, innate and cellular immunity, but was negatively associated with viral transcription, protein translation, humoral immunity, phagocytosis and complement activation [23]. In addition, besides the surface-bound ACE2, there exists another form of ACE2 that circulates freely in the blood stream [19]. It has been shown that soluble ACE2 can bind coronaviruses, competing with the surface-bound ACE2, thus preventing viral particle to bind to the cell [24]. Studies suggest that the activity of soluble ACE2 is higher in IBD [25]. This might contribute as a protective factor for IBD patients.

CD209L encoded by CLEC4M is a dendritic cell-specific ICAM-3-grabbing nonintegrin-related protein that has been described as an alternate receptor and portal of entry for severe acute respiratory syndrome coronavirus (SARS-CoV) [26]. Data showed that CD209L can mediate infection by SARS-CoV, although to a lesser extent than ACE2. CD209L is expressed on type II alveolar cells in human lung [26], but also in the intestine [27], both of which are main targets for coronavirus infection [28]. Ileal CLEC4M that encodes CD209L was revealed to have significantly decreased expression in Crohn’s disease (CD) [27] also suggesting possible protective implications against SARS-CoV-2 in CD patients.

Another molecular interaction between coronaviruses and IBD may be regarding endoplasmic reticulum (ER) stress. Pathogens as well as autoimmune and inflammatory diseases induce ER stress activating several signaling pathways [29]. The phosphorylation of the *α* subunit of eukaryotic translation initiation factor 2 (eIF2α) in response to ER stress stimulates the expression of viral proteins [30]. A fourfold elevation of the phosphorylated eIF2α was found in SARS-CoV infected cells [31]. On the contrary, eIF2α has an important role in mucosal homeostasis [32] and it has been reported that mechanisms induced by ER stress block phosphorylation of eIF2α in patients with ulcerative colitis [33].

#### Therapy challenges

It is uncertain whether SARS-CoV-2 induces severe forms of COVID-19 due to local viral replication or to subsequent immune system response [34]. The excessive and lengthened cytokine and chemokine response known as “*cytokine storm*” leads to high morbidity and mortality due to immunopathologic mechanisms. TNF plays pivotal roles in the “*cytokine storm*” immunopathology. TNF released by monocyte-macrophages boosts the apoptosis of lung epithelial and endothelial cells resulting in vascular leakage and alveolar edema that leads to hypoxia [35]. Also, TNF-mediated T cell apoptosis results in exuberant inflammatory reaction since T cells plays major role in subsiding hyperactive innate immune responses [36]. Additionally, coronavirus specific T cells have a key role in virus clearance [37]. In a study where TNF was the only neutralized pro-inflammatory cytokine, mice were protected from SARS-CoV-induced morbidity and mortality [38]. Thus, anti-TNF therapies in IBD might prove beneficial for patients in the context of COVID-19.

Another key cytokine produced in excess by activated macrophages is IL-6. A potential therapy targeting the host immune system in COVID-19 might be the cytokine blockade of IL-6 [39]. Tocilizumab, an anti-IL-6R antibody, also used in IBD treatment, is currently tested against COVID-19 in multiple clinical trials (NCT04317092 and NCT04346355).

One of the new drugs that is in clinical development for IBD, but already approved for the treatment of rheumatoid arthritis is Baricitinib, a selective janus kinase (JAK1, JAK2) and AP2-associated protein kinase 1 (AAK1) inhibitor. Baricitinib acts as both an anti-inflammatory drug through the inhibition of JAK1 and JAK2 and as an antiviral drug inhibiting receptor-mediated endocytosis by blocking AAK1 [22,40,41]. Currently, there are ongoing clinical trials for testing Baricitinib as a therapy for COVID-19 (NCT04346147).

Vedolizumab studies show that the rate of serious opportunistic infections and the frequency of tuberculosis were low and no hepatitis B/C viral reactivation was reported [42,22]. Studies showed similar opportunistic or viral infections in Ustekinumab therapy compared to placebo [22,43]. Blocking interferon signaling protected mice from lethal SARS-CoV infection and interferon secretion inhibition by Tofacitinib as shown in some studies [38], Tofacitinib used in IBD may be protective also against COVID-19.

Mesalazine (an antiinflammatory drug) and mercaptopurine (a thiopurine) used in IBD therapy were identified as putative repurposable drugs for potential treatment of SARS-CoV-2 through genomics and proteomics analysis using bioinformatics tools [44]. Indeed, studies shown that thiopurines were able to inhibit in vitro the papain-like protease of SARS-CoV and Middle East respiratory syndrome coronavirus that represents an important antiviral target essential in viral maturation and in the antagonism of interferon stimulation [45,46]. However, care must be taken in evaluating mesalazine as potential drug for COVID-19 since clinical studies showed possible pulmonary toxicities associations [47].

## 4. Conclusions

Certain insights into common molecular and therapy pathways regarding IBD patients infected with SARS-CoV-2 and other pathogenic coronaviruses, were described. Physicians need to make clinical decisions concerning the most suitable treatment management in IBD patients requiring immunosuppressive medication. The responsibility lies in correctly balancing infection risk with the risk of IBD relapse in case of treatment adjustment or discontinuation.

Using current understanding of SARS-CoV-2 as well as other pathogenic coronaviruses immunopathology, we showed why IBD patients might not be at an increased risk of infection or more severe outcomes. However, COVID-19 is a novel disease with possible different mechanisms of action as other related pathogens, with research still ongoing. Whether our findings are entirely applicable to the pathogenesis, disease susceptibility and treatment management of SARS-CoV-2 infection in IBD must be further explored.

## Data Availability

Data used to support the findings of this study are available from the corresponding author upon request.

## References

1. Organisation WH (2019) Coronavirus disease (COVID-19) Pandemic. https://www.who.int/emergencies/diseases/novel-coronavirus-2019. Accessed April 11th 2020

2. Mao R, Liang J, Shen J et al. Implications of COVID-19 for patients with pre-existing digestive diseases. Lancet Gastroenterol Hepatol 2020. 5 (5):426–428. doi:10.1016/s2468-1253(20)30076-5

3. Wen Z, Fiocchi C Inflammatory bowel disease: autoimmune or immune-mediated pathogenesis? Clin Dev Immunol 2004. 11 (3–4):195–204. doi:10.1080/17402520400004201

4. Xiao F, Tang M, Zheng X, Liu Y, Li X, Shan H Evidence for Gastrointestinal Infection of SARS-CoV-2. Gastroenterology 2020. doi:10.1053/j.gastro.2020.02.055

5. Kemp R, Dunn E, Schultz M Immunomodulators in Inflammatory Bowel Disease: An Emerging Role for Biologic Agents. BioDrugs 2013. 27 (6):585–590. doi:10.1007/s40259-013-0045-2

6. Long MD, Martin C, Sandler RS, Kappelman MD Increased Risk of Pneumonia Among Patients With Inflammatory Bowel Disease. American Journal of Gastroenterology 2013. 108 (2):240–248. doi:10.1038/ajg.2012.406

7. Tinsley A, Navabi S, Williams ED et al. Increased Risk of Influenza and Influenza-Related Complications Among 140,480 Patients With Inflammatory Bowel Disease. Inflammatory Bowel Diseases 2018. 25 (2):369–376. doi:10.1093/ibd/izy243

8. Click B, Regueiro M Managing Risks with Biologics. Current Gastroenterology Reports 2019. 21 (1):1. doi:10.1007/s11894-019-0669-6

9. Triantafillidis JK, Merikas E, Georgopoulos F Current and emerging drugs for the treatment of inflammatory bowel disease. Drug design, development and therapy 2011. 5: 185

10. Toruner M, Loftus EV, Jr., Harmsen WS et al. Risk Factors for Opportunistic Infections in Patients With Inflammatory Bowel Disease. Gastroenterology 2008. 134 (4):929–936. doi:10.1053/j.gastro.2008.01.012

11. Naganuma M, Kunisaki R, Yoshimura N, Takeuchi Y, Watanabe M A prospective analysis of the incidence of and risk factors for opportunistic infections in patients with inflammatory bowel disease. Journal of Gastroenterology 2013. 48 (5):595–600. doi:10.1007/s00535-012-0686-9

12. Ford AC, Peyrin-Biroulet L Opportunistic Infections With Anti-Tumor Necrosis Factor-α Therapy in Inflammatory Bowel Disease: Meta-Analysis of Randomized Controlled Trials. American Journal of Gastroenterology 2013. 108 (8):1268–1276. doi:10.1038/ajg.2013.138

13. Wang X, Zhou F, Zhao J et al. Elevated risk of opportunistic viral infection in patients with Crohn’s disease during biological therapies: a meta analysis of randomized controlled trials. European Journal of Clinical Pharmacology 2013. 69 (11):1891–1899. doi:10.1007/s00228-013-1559-8

14. Zingone F, Savarino EV Viral screening before initiation of biologics in patients with inflammatory bowel disease during the COVID-19 outbreak. The Lancet Gastroenterology & Hepatology. doi:10.1016/S2468-1253(20)30085-6

15. Pan L, Mu M, Yang P et al. Clinical Characteristics of COVID-19 Patients With Digestive Symptoms in Hubei, China: A Descriptive, Cross-Sectional, Multicenter Study. Am J Gastroenterol 2020. doi:10.14309/ajg.0000000000000620

16. Carvalho A, Alqusairi R, Adams A, Paul M, Kothari N, Peters S SARS-CoV-2 Gastrointestinal Infection Causing Hemorrhagic Colitis: Implications for Detection and Transmission of COVID-19 Disease. Am J Gastroenterol 2020.

17. Harmer D, Gilbert M, Borman R, Clark KL Quantitative mRNA expression profiling of ACE 2, a novel homologue of angiotensin converting enzyme. FEBS Letters 2002. 532 (1–2):107–110. doi:10.1016/s0014-5793(02)03640-2

18. Rao S, Lau A, So H-C Exploring diseases/traits and blood proteins causally related to expression of ACE2, the putative receptor of 2019-nCov: A Mendelian Randomization analysis. medRxiv 2020.2020.2003.2004.20031237. doi:10.1101/2020.03.04.20031237

19. Monteleone G, Ardizzone S Are Patients with Inflammatory Bowel Disease at Increased Risk for Covid-19 Infection? Journal of Crohn’s and Colitis 2020. doi:10.1093/ecco-jcc/jjaa061

20. Jablaoui A, Kriaa A, Mkaouar H et al. Fecal Serine Protease Profiling in Inflammatory Bowel Diseases. Front Cell Infect Microbiol 2020. 10:21. doi:10.3389/fcimb.2020.00021

21. Gastroenterology ACo (2020) COVID-19 and GI. https://gi.org/media/covid-19-and-gi/. Accessed April 11th 2020

22. Rubin DT, Abreu MT, Rai V, Siegel CA Management of Patients with Crohn’s Disease and Ulcerative Colitis During the COVID-19 Pandemic: Results of an International Meeting. Gastroenterology. doi:10.1053/j.gastro.2020.04.002

23. Wang J, Zhao S, Liu M et al. ACE2 expression by colonic epithelial cells is associated with viral infection, immunity and energy metabolism. medRxiv 2020.2020.2002.2005.20020545. doi:10.1101/2020.02.05.20020545

24. Batlle D, Wysocki J, Satchell K Soluble angiotensin-converting enzyme 2: a potential approach for coronavirus infection therapy? Clinical Science 2020. 134 (5):543–545. doi:10.1042/cs20200163

25. Garg M, Burrell LM, Velkoska E et al. Upregulation of circulating components of the alternative reninangiotensin system in inflammatory bowel disease: A pilot study. Journal of the Renin-Angiotensin-Aldosterone System 2015. 16 (3):559–569. doi:10.1177/1470320314521086

26. Jeffers SA, Tusell SM, Gillim-Ross L et al. CD209L (L-SIGN) is a receptor for severe acute respiratory syndrome coronavirus. Proceedings of the National Academy of Sciences of the United States of America 2004. 101 (44):15748–15753. doi:10.1073/pnas.0403812101

27. Hughes AL Consistent across-tissue signatures of differential gene expression in Crohn’s disease. Immunogenetics 2005. 57 (10):709–716. doi:10.1007/s00251-005-0044-7

28. Gu J, Korteweg C Pathology and Pathogenesis of Severe Acute Respiratory Syndrome. The American Journal of Pathology 2007. 170 (4):1136–1147. doi:10.2353/ajpath.2007.061088

29. Smith JA Regulation of Cytokine Production by the Unfolded Protein Response; Implications for Infection and Autoimmunity. Frontiers in Immunology 2018. 9 (422). doi:10.3389/fimmu.2018.00422

30. He B Viruses, endoplasmic reticulum stress, and interferon responses. Cell Death & Differentiation 2006. 13 (3):393–403. doi:10.1038/sj.cdd.4401833

31. Chan C-P, Siu K-L, Chin K-T, Yuen K-Y, Zheng B, Jin D-Y Modulation of the Unfolded Protein Response by the Severe Acute Respiratory Syndrome Coronavirus Spike Protein. Journal of Virology 2006. 80 (18):9279–9287. doi:10.1128/jvi.00659-06

32. Luo K, Cao SS Endoplasmic reticulum stress in intestinal epithelial cell function and inflammatory bowel disease. Gastroenterol Res Pract 2015. 2015:328791. doi:10.1155/2015/328791

33. Tréton X, Pédruzzi E, Cazals–Hatem D et al. Altered Endoplasmic Reticulum Stress Affects Translation in Inactive Colon Tissue From Patients With Ulcerative Colitis. Gastroenterology 2011. 141 (3):10241035. doi:https://doi.org/10.1053/j.gastro.2011.05.033

34. Ledford H How does COVID-19 kill? Uncertainty is hampering doctors’ ability to choose treatments. Nature 2020. 580 (7803):311–312. doi:10.1038/d41586-020-01056-7

35. Channappanavar R, Perlman S Pathogenic human coronavirus infections: causes and consequences of cytokine storm and immunopathology. Seminars in Immunopathology 2017. 39 (5):529–539. doi:10.1007/s00281-017-0629-x

36. Dong Kim K, Zhao J, Auh S et al. Adaptive immune cells temper initial innate responses. Nature Medicine 2007. 13 (10):1248–1252. doi:10.1038/nm1633

37. Zhao J, Zhao J, Legge K, Perlman S Age-related increases in PGD2 expression impair respiratory DC migration, resulting in diminished T cell responses upon respiratory virus infection in mice. The Journal of Clinical Investigation 2011. 121 (12):4921–4930. doi:10.1172/JCI59777

38. Channappanavar R, Fehr Anthony R, Vijay R et al. Dysregulated Type I Interferon and Inflammatory Monocyte-Macrophage Responses Cause Lethal Pneumonia in SARS-CoV-Infected Mice. Cell Host & Microbe 2016. 19 (2):181–193. doi:10.1016/j.chom.2016.01.007

39. Liu B, Li M, Zhou Z, Guan X, Xiang Y Can we use interleukin-6 (IL-6) blockade for coronavirus disease 2019 (COVID-19)-induced cytokine release syndrome (CRS)? Journal of Autoimmunity 2020.102452. doi:https://doi.org/10.1016/j.jaut.2020.102452

40. Favalli EG, Biggioggero M, Maioli G, Caporali R Baricitinib for COVID-19: a suitable treatment? Lancet Infect Dis 2020. doi:10.1016/s1473-3099(20)30262-0

41. Richardson P, Griffin I, Tucker C et al. Baricitinib as potential treatment for 2019-nCoV acute respiratory disease. Lancet 2020. 395 (10223):e30-e31. doi:10.1016/s0140-6736(20)30304-4

42. Ng SC, Hilmi IN, Blake A et al. Low Frequency of Opportunistic Infections in Patients Receiving Vedolizumab in Clinical Trials and Post-Marketing Setting. Inflammatory Bowel Diseases 2018. 24 (11):2431–2441. doi:10.1093/ibd/izy153

43. Sandborn WJ, Feagan BG, Fedorak RN et al. A Randomized Trial of Ustekinumab, a Human Interleukin-12/23 Monoclonal Antibody, in Patients With Moderate-to-Severe Crohn’s Disease. Gastroenterology 2008. 135 (4):1130–1141. doi:10.1053/j.gastro.2008.07.014

44. Zhou Y, Hou Y, Shen J, Huang Y, Martin W, Cheng F Network-based drug repurposing for novel coronavirus 2019-nCoV/SARS-CoV-2. Cell Discovery 2020. 6 (1):14. doi:10.1038/s41421-020-0153-3

45. Chou C-Y, Chien C-H, Han Y-S et al. Thiopurine analogues inhibit papain-like protease of severe acute respiratory syndrome coronavirus. Biochemical Pharmacology 2008. 75 (8):1601–1609. doi:https://doi.org/10.1016/j.bcp.2008.01.005

46. Cheng K-W, Cheng S-C, Chen W-Y et al. Thiopurine analogs and mycophenolic acid synergistically inhibit the papain-like protease of Middle East respiratory syndrome coronavirus. Antiviral Research 2015. 115:9–16. doi:https://doi.org/10.1016/j.antiviral.2014.12.011

47. Gupta A, Gulati S Mesalamine induced eosinophilic pneumonia. Respiratory Medicine Case Reports 2017. 21:116–117. doi:https://doi.org/10.1016/j.rmcr.2017.04.010

